# Prevalence of central obesity and its associated risk factors among adults in Southeast Ethiopia: a community-based cross-sectional study

**DOI:** 10.1101/2022.02.24.22271452

**Authors:** Yohannes Tekalegn, Damtew Solomon, Biniyam Sahiledengle, Tesfaye Assefa, Wegene Negash, Anwar Tahir, Tadele Regassa, Ayele Mamo, Habtamu Gezahegn, Kebebe Bekele, Demisu Zenbaba, Alelign Tasew, Fikreab Desta, Daniel Atilaw, Zegeye Regassa, Fikadu Nugusu, Zinash Teferu Engida, Degefa Gomora Tesfaye, Chala Kene, Wondu Shiferaw Nigussie, Dereje Chala, Adisu Gemechu Abdi, Girma Beressa, Demelash Woldeyohannes, Heather L. Rogers, Lillian Mwanri

## Abstract

**Background:** Obesity and overweight are known are public health scourge challenges affecting populations across the world. These conditions have been associated with a wide range of chronic diseases including type 2 diabetes mellitus, cardiovascular disease, and cancers. In Ethiopia, literature regarding the burden of central (abdominal) obesity is scarce. This study aimed to fill this gap by assessing the prevalence and risk factors associated with central obesity among adults in Ethiopia.

**Methods:** From May to July 2021, a community-based cross-sectional survey was conducted on a sample of 694 adults aged ≥18 years in administrative towns of Bale zone, Southeast Ethiopia. Multi-stage sampling followed by systematic random sampling was employed to identify study participants. Waist and hip circumferences were measured using standard protocols. The World Health Organization STEPS wise tool was used to assess risk factors associated with central obesity. Bi-variable and multi-variable binary logistic regression were used to identify factors associated with central obesity. Adjusted odds ratios (AOR) and their corresponding 95% confidence intervals (CI) were reported to estimate the strength of associations.

**Results:** The overall prevalence of central obesity using waist circumference was 39.01% (15.44% for men and 53.12% for women). Multi-variable binary logistic regression analysis revealed that female sex (AOR=12.93, 95% CI: 6.74-24.79), Age groups: 30-39 years old (AOR=2.8, 95% CI: 1.59-4.94), 40-49 years (AOR=7.66, 95% CI: 3.87-15.15), 50-59 years (AOR=4.65, 95% CI: 2.19-9.89), ≥60 years (AOR=12.67, 95% CI: 5.46-29.39), occupational status like: housewives (AOR=5.21, 95% CI: 1.85-14.62), self-employed workers (AOR=4.63, 95% CI: 1.62-13.24), government/private/non-government employees (AOR=4.68, 95% CI: 1.47-14.88) and skipping breakfast (AOR=0.46, 95% CI: 0.23-0.9) were significantly associated with central obesity.

**Conclusions:** Abdominal obesity has become an epidemic in towns of southeast Ethiopia, and the prevalence is higher among women. Female sex, older age group, being employed, not skipping breakfast were significantly associated with central obesity.

## Introduction

The World Health Organization (WHO) defines obesity as abnormal or excess fat accumulation that may impair health (1). Obesity has been associated with the imbalance between intake and energy expenditure, a condition that may be caused by either non-modifiable or modifiable factors (2). This caloric imbalance creates an excess accumulation of energy which in turn is stored in the body resulting in excess body weight. Although genetic factors are not modifiable, obesity can result from a complex interaction between environmental, socio-economic, and/or personal behaviors, factors that are modifiable. Addressing the modifiable factors play as one of the critical strategies in preventing obesity (3). Globally, the prevalence of overweight and obesity increased by 27.5% for adults and 47.1% for children between 1980 and 2013. The number of individuals with overweight and obesity increased from 857 million in 1980 to 2.1 billion in 2013 (4). If this secular trend continues persistently, 38% of the world’s adult population will be overweight and another 20% will be obese by 2030 (5).

Worldwide, a high body mass index (BMI) has been reported to be responsible for 4 million deaths and 120 million disability-adjusted life years (6). Several studies have further documented obesity as the risk factor for many non-communicable diseases (NCDs) and chronic health conditions including hypertension, high lipid concentrations, type 2 diabetes, coronary heart disease, stroke, and certain cancers (7-22). Central (abdominal) obesity, measured in waist circumference, waist to hip ratio, and waist to height ratios, is highly linked with increased risk of morbidity and mortality and is considered to be superior to BMI in predicting cardiovascular disease and mortality risks (23-25).

Many low and middle-income countries (LMICs), including Ethiopia, currently face a double burden of malnutrition (26). While LMICs are dealing with problems of infectious diseases and undernutrition, they are also experiencing a rapid increase in non-communicable disease risk factors like overweight and obesity (1, 27-29), and the emergence of Corona Virus Disease (COVID-19) pandemic has further challenged health systems, economies and populations across the globe, but and more severely in LMICs (30, 31). Evidence suggests that NCDs are dramatically increasing in Ethiopia and it was estimated that NCDs were responsible for 711 deaths per 100,000 population in 2015 (32). Cardiovascular diseases, cancer, diabetes, and mental disorders were responsible for 30% of the total disease burden in Ethiopia as measured in age-standardized disability-adjusted life years (DALYs) rates in 2017 (33).

In Ethiopia, epidemiological studies regarding the prevalence, distribution, and determinants of obesity are meager. More specifically, few studies have assessed the prevalence and risk factors of central (abdominal) obesity (34-39), a superior predictor of NCDs. The prevalence reported in the aforementioned previous studies ranges from 15.5% in the northern part of Ethiopia (36) to 37.4% in southwest Ethiopia (39). Similarly, risk factors identified by those studies vary from district to district which warrants studying the prevalence and context-based risk factors in our study setting which has implications for prevention efforts / public health campaigns. To the best of our knowledge, there is no published evidence on the magnitude and factors associated with central obesity in Southeast Ethiopia. The current study aims to assess the prevalence of central (abdominal) obesity and its associated risk factors in the administrative towns of Bale Zone, Southeast Ethiopia.

## Methods and Materials

### Study design, setting and subjects

From May to July 2021, a community-based cross-sectional study was conducted to assess the prevalence of central (abdominal) obesity among adults (≥18 years) in the administrative towns of Bale zone, Southeast Ethiopia. All adults residing in the study area for at least 6 months were eligible for inclusion. However, potential participants were excluded if they had: psychiatric problems, hearing impairments, body deformities (kyphosis and scoliosis), other debilitations and/ or handicaps. Pregnant women were also ineligible for inclusion. Bale zone, one of the Regional States in Ethiopia in the Southeastern part of Oromia (an area of 43,690.56 km^2^), is located between 5^0^ 22’ – 8^0^ 08’ latitude north and between 38^0^ 41’-40^0^ 44’ longitude east. **(Figure 1)**. According to the Central Statistical Agency (CSA), in 2007 (latest available report), the Bale zone, had a total population of 1,402,492, including 713,517 men and 688,975 women, an increase of 15.16% from the 1994 Census. Of the total inhabitants in Bale Zone, 166,758 (26.20% were) urban dwellers and 44,610 (3.18% were) pastoralists. The zone comprises 18 districts and a total of 297,081 (an average of 4.72 persons per household) households. There are two administrative towns in the zone namely Robe and Goba towns. Furthermore, each administrative town is divided into smaller administrative clusters known as ***‘****gots’*. Robe and Goba towns have 36 and 24 *gots* respectively. According to the 2021 administrative report, Robe and Goba towns respectively had population of 73,152 and 52,785 people.

### Sample size determination and sampling procedures

Based on the total population in the zone, the study sample size of 700 individuals was calculated using OpenEpi (Version 3, an open-source calculator)considering the following parameters: 95% level of confidence, 4% margin of error, 24.4% reported prevalence of abdominal obesity by the previous study in Dilla town, South Ethiopia (38), design effect of 2, and non-response rate of 5%.. A multi-stage stratified sampling and, systematic random sampling were employed to select the study participants for inclusion in the study. Initially, the study populations sampling was stratified into the two (Robe and Goba) towns, followed by stratification into randomly selected clusters (*gots*). One third of clusters were selected from each town, and proportionate to their number, eight and 12 *gots* were selected from Goba and Robe respectively. Furthermore, households in the sampled clusters were selected using systematic sampling techniques and one adult per sampled household was selected using the lottery method.

### Data collection and measurement procedures

Interviewer-administered, structured questionnaires were used to collect data on socio-demographic and behavioural characteristics followed by physical measurements of weight, height, waist, and hip circumferences. A standard questionnaire was adapted from the WHO STEPS-wise questionnaire for chronic disease risk factor surveillance (40). The English version of the questionnaire was translated to the local languages spoken in the study area, Afan Oromo and Amharic. After the translation into local languages, the questionnaire was back-translated to English to check the consistency. Weight was measured using an electronic digital weight scale by putting the scale on a firm flat surface after participants took off footwear, heavy clothes, and empty their pockets for heavy items. Participants’ height measurements were taken in a standing position by a portable height measuring board placed on a firm surface against a wall. With participants facing the data collector, feet placed together, and eyes leveled at the ears, readings were taken in centimeters (to the nearest millimeter). With the participants’ arms relaxed at the sides, the waist circumferences were measured by constant tension tape at the end of a normal expiration; at the midpoint between the lower margin of the last palpable rib and the top of the iliac crest (hip bone). Waist measurements read at the level of the tape to the nearest millimeter, while ensuring the tape was comfortably tight enough not to cause compression of the skin. Hip circumferences were measured by constant tension tape with the arms relaxed at the sides at the maximum circumference over the buttocks. Hip circumferences were measured and read at the level of the tape to the nearest millimeter. Further details on the study physical measurement protocols are consistent with the WHO STEPS-wise instrument guideline (41).

Data collection was conducted by six data collectors (three male-female pairs) with degrees of bachelor in health sciences (Nursing, Public Health, and Midwifery) Female and male data collectors respectively collected female and male participants’ data. Two supervisors with a degree master of public health took part in overseeing the data collection process. Data collectors and supervisors were provided with a two-day intensive training on the objective of the study, administration protocols for the questionnaires, administration protocols for anthropometric measurements (weight, height, waist, and hip circumferences), and how to maintain confidentiality and privacy of the study participants. With assistance from the study supervisors, all questionnaires were checked every day by data collectors for completeness before leaving the data collection site (household).

### Outcome variable

Central (abdominal) obesity is the dependent variable for this study, which is derived from waist circumference measurement. As per World Health Organization recommendations, waist-circumference >94 centimeters for males and >80 centimeters for females were categorized as central obesity (23). Individuals with central obesity were coded as “1” and others were coded as “0”.

### Independent variables

Independent variables included sociodemographic and behavioral variables. Sociodemographic variables included the town of residence (Robe or Goba); age (categorized into 18-29, 30-39, 40-49, 50-59, and ≥ 60 years); sex (male or female), marital status (categorized as never married, married/cohabiting, or divorced/separated/widowed); educational status (categorized as no formal education, secondary education, or diploma and above); occupational status (housewives, self-employed, government/non-government/private employee, student/unemployed, or Retired); family size (categorized as ≤ 2, >2); and wealth index (computed by principal component analysis using household assets and rank-ordered as low, medium or high wealth index terciles). Behavioral variables included characteristics such as fruit and vegetable consumption (categorized as less than five servings of fruit and/or vegetables per day, or five or more servings of fruit and/or vegetables per day), skipping breakfast (categorized as yes or no), avoidance of eating foods prepared outside the home (categorized as yes or no) and levels of physical activities [derived by calculating the metabolic equivalent value (MET) - study participants with an equivalent combination of moderate and vigorous-intensity physical activity achieving at least 600 MET-minutes were categorized as sufficient level while the others were categorized as insufficient physical activity levels], sedentary activities were measure by adding total time spent sitting or reclining on a typical day, current smokers of tobacco were categorized as yes or no and participants who consumed alcohol were categorized as never drank, consumed in last 12 months, or consumed in last 30 days. Measurements including fruit and/or vegetable consumption, level of physical activities, alcohol, and/or tobacco consumption were assessed and analyzed using World Health Organization (WHO) STEPS Surveillance tool recommendations (41).

### Data analysis procedures

The data was coded and entered into EpiData Version 3.1 and cleaned, processed, and analyzed using SPSS version 25 and STATA version 14. The variables were described using mean, frequencies, proportions, and tables. The Chi-square test was used to check the statistical difference of the distribution of categorical independent variables between men and women. A two-sample Wilcoxon rank-sum (Mann-Whitney) test was used to check the statistical difference in the distribution of continuous independent variables between men and women. Both bi-variable and multi-variable binary logistic regression analyses were used to identify factors associated with the outcome variable. Variables having a p-value of less than 0.25 in the bi-variable binary logistic regression model were included in the multivariable binary logistic regression analysis model. After checking for multi-collinearity, enter method was used to run the model. The logit of the dependent variable was checked for outlier and 7 outlying values (having standardized residual >2.58 at the level of α<0.01) were excluded from the analysis. Hosmer and Lemeshow’s goodness of model fit was checked and the data fitted the model well (p=0.92). Multi-collinearity between independent variables was checked using variance inflation factor (VIF), the mean VIF was 2.1 which is less than the recommended cut-off values (42). Finally, adjusted odds ratios with 95% confidence intervals were used to estimate the strength of associations between the outcome variable and independent variables. All tests were two-tailed and statistical significances were declared at p-value <0.05.

### Ethical considerations

Ethical clearance and support letters to introduce the researchers and the study to respective study areas were obtained from the ethical review committee of Madda Walabu University. Permission letters to conduct the survey were obtained from the respective authorities of the two towns (Goba and Robe). The methods were conducted following the tenets of the Helsinki declaration. To obtain oral informed consent an information sheet was read to all eligible study participants before data were collected. The privacy of the respondents was respected and data were de-identified before analysis and were reported in aggregate.

## Results

### Sociodemographic characteristics of study participants

A total of 694 adults (259 men and 435 women) participated in this study, a response rate of 99.1%. Four hundred and eight (58.8%) and 286 (41.2%) study participants were from Robe and Goba town respectively. Participants’ age ranged from 18-95 years old. Men participants’ median age was 41 years old with an interquartile range (IQR) of 28-55 years. women’s median age was 32 years with an IQR of 25-45 years (**Table 1**).

**Table 1:**
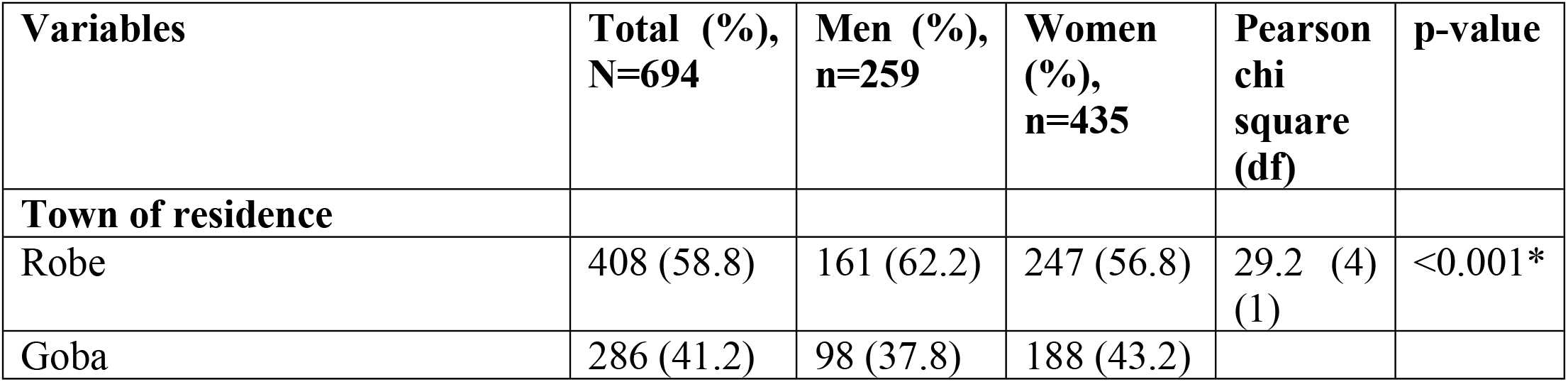
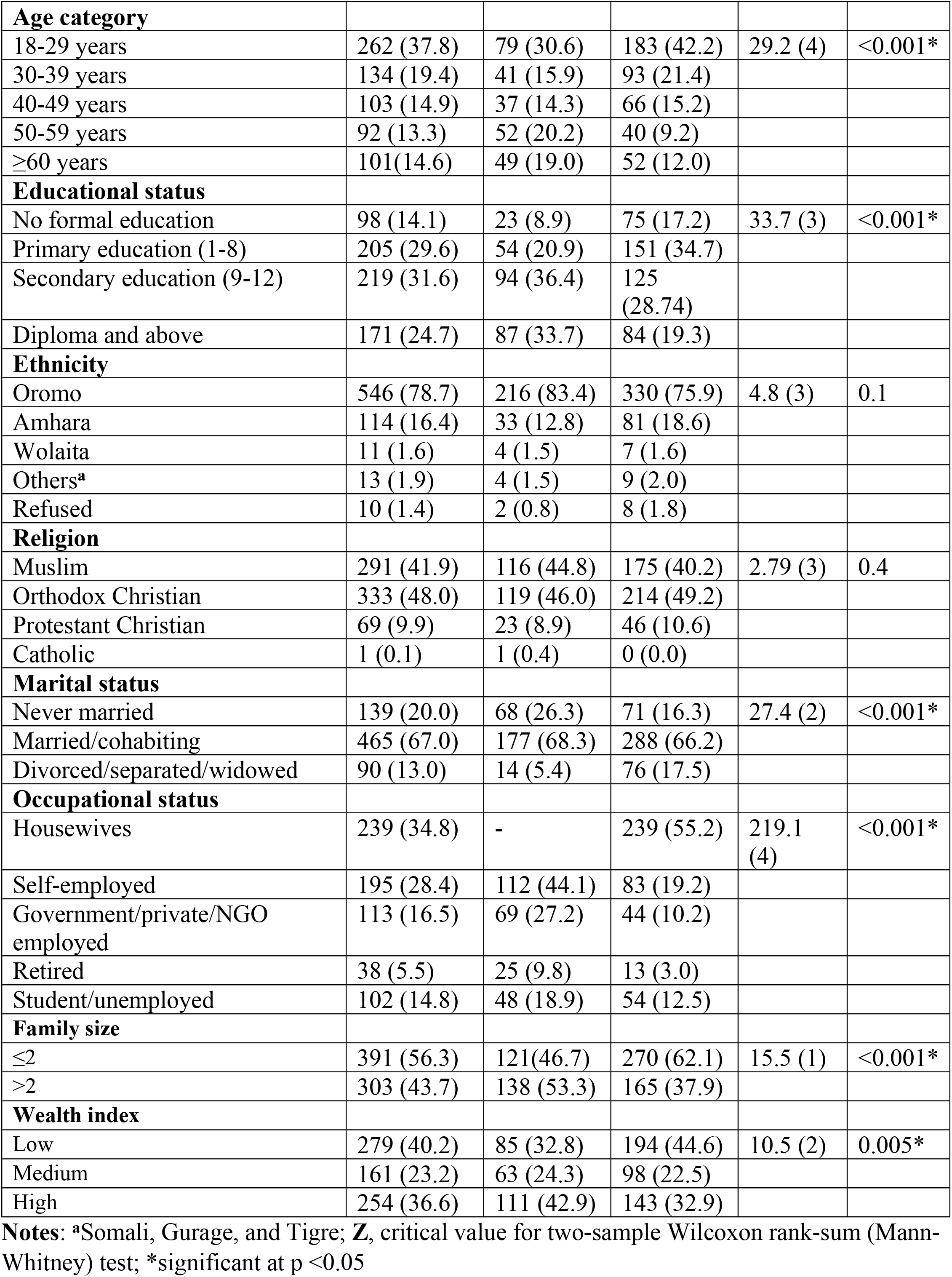
Sociodemographic characteristics of study participants in administrative towns of Bale Zone, Southeast Ethiopia, 2021.

| Variables | Total (%),<br>N=694 | Men (%),<br>n=259 | Women<br>(%),<br>n=435 | Pearson<br>chi<br>square<br>(df) | p-value |
| --- | --- | --- | --- | --- | --- |
| **Town of residence** |  |  |  |  |  |
| Robe | 408 (58.8) | 161 (62.2) | 247 (56.8) | 29.2 (4)<br>(1) | <0.001* |
| Goba | 286 (41.2) | 98 (37.8) | 188 (43.2) |  |  |
| **Age category** |  |  |  |  |  |
| 18-29 years | 262 (37.8) | 79 (30.6) | 183 (42.2) | 29.2 (4) | <0.001* |
| 30-39 years | 134 (19.4) | 41 (15.9) | 93 (21.4) |  |  |
| 40-49 years | 103 (14.9) | 37 (14.3) | 66 (15.2) |  |  |
| 50-59 years | 92 (13.3) | 52 (20.2) | 40 (9.2) |  |  |
| ≥60 years | 101(14.6) | 49 (19.0) | 52 (12.0) |  |  |
| **Educational status** |  |  |  |  |  |
| No formal education | 98 (14.1) | 23 (8.9) | 75 (17.2) | 33.7 (3) | <0.001* |
| Primary education (1-8) | 205 (29.6) | 54 (20.9) | 151 (34.7) |  |  |
| Secondary education (9-12) | 219 (31.6) | 94 (36.4) | 125 (28.74) |  |  |
| Diploma and above | 171 (24.7) | 87 (33.7) | 84 (19.3) |  |  |
| **Ethnicity** |  |  |  |  |  |
| Oromo | 546 (78.7) | 216 (83.4) | 330 (75.9) | 4.8 (3) | 0.1 |
| Amhara | 114 (16.4) | 33 (12.8) | 81 (18.6) |  |  |
| Wolaita | 11 (1.6) | 4 (1.5) | 7 (1.6) |  |  |
| Others<sup>a</sup> | 13 (1.9) | 4 (1.5) | 9 (2.0) |  |  |
| Refused | 10 (1.4) | 2 (0.8) | 8 (1.8) |  |  |
| **Religion** |  |  |  |  |  |
| Muslim | 291 (41.9) | 116 (44.8) | 175 (40.2) | 2.79 (3) | 0.4 |
| Orthodox Christian | 333 (48.0) | 119 (46.0) | 214 (49.2) |  |  |
| Protestant Christian | 69 (9.9) | 23 (8.9) | 46 (10.6) |  |  |
| Catholic | 1 (0.1) | 1 (0.4) | 0 (0.0) |  |  |
| **Marital status** |  |  |  |  |  |
| Never married | 139 (20.0) | 68 (26.3) | 71 (16.3) | 27.4 (2) | <0.001* |
| Married/cohabiting | 465 (67.0) | 177 (68.3) | 288 (66.2) |  |  |
| Divorced/separated/widowed | 90 (13.0) | 14 (5.4) | 76 (17.5) |  |  |
| **Occupational status** |  |  |  |  |  |
| Housewives | 239 (34.8) | - | 239 (55.2) | 219.1 (4) | <0.001* |
| Self-employed | 195 (28.4) | 112 (44.1) | 83 (19.2) |  |  |
| Government/private/NGO employed | 113 (16.5) | 69 (27.2) | 44 (10.2) |  |  |
| Retired | 38 (5.5) | 25 (9.8) | 13 (3.0) |  |  |
| Student/unemployed | 102 (14.8) | 48 (18.9) | 54 (12.5) |  |  |
| **Family size** |  |  |  |  |  |
| ≤2 | 391 (56.3) | 121(46.7) | 270 (62.1) | 15.5 (1) | <0.001* |
| >2 | 303 (43.7) | 138 (53.3) | 165 (37.9) |  |  |
| **Wealth index** |  |  |  |  |  |
| Low | 279 (40.2) | 85 (32.8) | 194 (44.6) | 10.5 (2) | 0.005* |
| Medium | 161 (23.2) | 63 (24.3) | 98 (22.5) |  |  |
| High | 254 (36.6) | 111 (42.9) | 143 (32.9) |  |  |

**Notes:** <sup>a</sup>Somali, Gurage, and Tigre; **Z**, critical value for two-sample Wilcoxon rank-sum (Mann-Whitney) test; \*significant at p <0.05

### Behavioral characteristics of the study participants

More than half (58%) of the study participants consumed less than five servings of fruit and/or vegetable per day. The mean number of days of fruit were consumed per week was 2.3 days. Approximately 15% of the study participants reported skipping breakfast and another 15% avoided eating foods prepared outside the home. Nearly 29% of the study participants had an insufficient level of physical activity as per World Health Organization recommendations (**Table 2**).

**Table 2:** Behavioral characteristics of study participants in administrative towns of Bale zone, Southeast Ethiopia, 2021.

| Variables | Total (%),<br>N=694 | Men (%),<br>n=259 | Women<br>(%),<br>n=435 | Pearson<br>chi<br>square<br>(df) | p-value |
| --- | --- | --- | --- | --- | --- |
| **Number of days eat fruit and/or vegetables on average per week** |  |  |  |  |  |
| No fruit and/or vegetable | 58 (8.4) | 30 | 28 (6.4) | 77 (3) | 0.05 |
| 1-2 | 349 (50.4) | 118 | 231 (53.1) |  |  |
| 3-4 | 219 (31.6) | 87 | 132 (30.3) |  |  |
| >=5 | 67 (9.7) | 23 | 44 (10.1) |  |  |
| **Number of servings of fruit and/or vegetables on average per day** |  |  |  |  |  |
| 1-2 servings | 49 (8.1) | 17 (8.0) | 32 (8.2) | 0.4 (2) | 0.8 |
| 3-4 servings | 301 (49.8) | 102 (48.3) | 199 (50.9) |  |  |
| >=5 servings | 252 (41.7) | 92 (43.6) | 160 (40.9) |  |  |
| **Avoid eating foods prepared outside of a home** |  |  |  |  |  |
| Yes | 584 (84.6) | 176 (68.2) | 408 (94.4) | 85.4 (1) | <0.001* |
| No | 106 (15.4) | 82 (31.8) | 24 (5.6) |  |  |
| **Skip breakfast** |  |  |  |  |  |
| No | 592 (14.5) | 210 (81.4) | 382 (88.0) | 5.7 (1) | 0.01* |
| Yes | 100 (88.5) | 48 (18.6) | 52 (12.0) |  |  |
| **Level of physical activity** |  |  |  |  |  |
| Insufficient physical activity | 195 (28.8) | 31 (12.3) | 164 (38.6) | 53.3 (1) | <0.001* |
| Sufficient physical activity | 482 (71.2) | 221 (87.7) | 261 (61.4) |  |  |
| **Number of days fruit consumed in a typical week ( mean ± SD)** | 2.33 ± 1.60 | 2.31 ± 1.57 | 2.35 ± 1.62 | Z= 0.1 | 0.9 |
| **Number of days vegetables consumed in a typical week ( mean ± SD)** | 3.59 ± 1.97 | 3.49 ± 1.83 | 3.66 ± 2.05 | Z = -0.45 | 0.6 |
| **Number of servings of fruit on average per day ( mean ± SD)** | 2.11 ± 0.86 | 2.25 ± 0.99 | 2.04 ± 0.77 | Z = 2.28 | 0.02* |
| **Number of servings of vegetables on average per day ( mean ± SD)** | 2.11 ± 0.96 | 2.11 ± 0.98 | 2.12 ± 0.95 | Z = -0.28 | 0.7 |
| **Number of servings of fruit and/or vegetables on average per day ( mean ± SD)** | 4.2 ± 1.40 | 4.36 ± 1.54 | 4.14 ± 1.31 | Z = 1.35 | 0.1 |
| **Minutes spent on sedentary activities on average per day ( mean ± SD)** | 178.1 ± 128.98 | 166.65 ± 125.25 | 184.90 ± 130.80 | Z = -1.78 | 0.07 |

Notes: Z, critical value for two-sample Wilcoxon rank-sum (Mann-Whitney) test; \*significant at p < 0.05

### Alcohol and tobacco use behavioural characteristics of the study participants

Only 7 (1%) indicated currently smoking tobacco products. Two hundred twenty-two (32.0%) consumed alcohol in the past twelve months (**Table 3**).

**Table 3:** Alcohol and tobacco consumption behavior of study participants in administrative towns of Bale zone, Southeast Ethiopia, 2021.

| **Variables** | **Total**<br>**(%),**<br>**N=694** | **Men (%),**<br>**n=259** | **Women**<br>**(%),**<br>**n=435** | **Pearson**<br>**chi**<br>**square**<br>**(df)** | **p-value** |
| --- | --- | --- | --- | --- | --- |
| **Currently smoke tobacco products, n=694** |  |  |  |  |  |
| Yes | 7 (1.0) | 7 (2.7) | 0 (0.0) | 11.87<br>(1) | <0.001* |
| No | 687<br>(99.0) | 252<br>(97.3) | 435 (100) |  |  |
| **Ever consumed alcohol, n=693** |  |  |  |  |  |
| Yes | 222<br>(32.0) | 94 (36.4) | 128 (29.4) | 3.65 (1) | 0.05 |
| No | 471<br>(68.0) | 164<br>(63.6) | 307 (70.6) |  |  |
| **Consumed alcohol in past 12 months, n=222** |  |  |  |  |  |
| Yes | 212<br>(95.5) | 90 (94.7) | 122 (96.1) | 0.22 (1) | 0.6 |
| No | 10 (4.5) | 5 (5.3) | 5 (3.9) |  |  |
| **Frequency of alcohol drinking in past 12 months, n=222** |  |  |  |  |  |
| 5-6 days per week | 1 (0.5) | 0 (0.0) | 1 (0.8) | 38.50<br>(3) | <0.001* |
| 1-4 days per week | 25 (11.7) | 18 (20.0) | 7 (5.7) |  |  |
| 1-3 days per week | 89 (41.8) | 52 (57.8) | 37 (30.1) |  |  |
| Less than once a month | 97 (46.0) | 20 (22.2) | 78 (63.4) |  |  |
| **Consumed alcohol within past 30 days, n=222** |  |  |  |  |  |
| Yes | 134<br>(60.5) | 68 (71.6) | 67 (52.3) | 8.44 (1) | 0.004* |
| no | 88 (39.5) | 27 (28.4) | 61 (47.7) |  |  |
| **Five or more drinks on a single occasion at least once during the past 30 days, n=135** |  |  |  |  |  |
| Yes | 23 (17.0) | 15 (5.8) | 8 (1.8) | 7.91 (1) | 0.005* |
| No | 112<br>(83.0) | 244<br>(94.2) | 427 (98.2) |  |  |
| **Number of drinking occasions in past 30 days among current (30 days) drinkers ( mean ± SD), n=134** | 2.53 ±<br>1.96 | 3.13 ±<br>2.14 | 1.92 ± 1.54 | Z = 4.63 | <0.001* |
| **Number of standard drinks per drinking occasion among current (past 30 days) drinkers ( mean ± SD), n=133** | 2.35 ±<br>1.20 | 2.73 ±<br>1.29 | 1.97 ± 0.98 | Z = 3.77 | <0.001* |
| **Maximum number of standard drinks consumed on one occasion in the past 30 days ( mean ± SD), n=134)** | 2.87 ±<br>.47 | 3.52 ±<br>1.63 | 2.24 ± 0.96 | Z = 5.33 | <0.001* |

Notes: Z, critical value for two-sample Wilcoxon rank-sum (Mann-Whitney) test; \*significant at p < 0.05

### Prevalence of central obesity

As described in **Table 4**, of the 259 men included in this study, 40 (15.44%, 95% CI: 11.26-20.43%) had waist circumference >94 centimeters. Of the 433 women included in the study, 230 (53.12%, 95% CI: 48.29-57.89%) had a waist circumference greater than 80 centimeters. Based on the waist-to-hip ratios, 204 (47.11%, 95% CI: 42.32, 51.93%) and 129 (49.80%, 95% CI: 43.55-56.06%) of women and men had wait to hip ratio of greater than or equal to 0.85 and 0.90, respectively.

**Table 4:** Distribution of central obesity by sex among adult populations in administrative towns of Bale zone, Southeast Ethiopia, 2021.

| Central obesity distribution by sex | Frequency | Percent (95% CI) |
| --- | --- | --- |
| **Men (n= 259)** |  |  |
| Waist circumference > 94 centimeters | 40 | 15.44 (11.26-20.43) |
| Waist circumference > 102 centimeters | 9 | 3.47 (1.60-6.49) |
| Waist to hip ratio ≥ 0.9 | 129 | 49.81 (43.55-56.06) |
| **Women (n=433 )** |  |  |
| Waist circumference > 80 centimeters | 230 | 53.12 (48.29-57.89) |
| Waist circumference > 88 centimeters | 118 | 27.19 (23.11-31.70) |
| Waist to hip ratio ≥ 0.85 | 204 | 47.11 (42.32-51.93) |
| **<sup>a</sup> Overall (both sexes) N=692** |  |  |
| Centrally obese | 270 | 39.01 (35.36-42.76) |

**Notes:** <sup>a</sup> calculated using waist circumference >94 and >80 centimeters for men and women respectively.

### Factors associated with central (abdominal) obesity

Findings of the multivariable binary logistic regression analysis revealed that women had higher odds of central obesity compared to men (AOR:12.93, 95% CI: 6.74-24.79). Age groups 30-39 (AOR: 2.80, 95% CI: 1.59-4.94), 40-49 (AOR: 7.66, 95% CI: 3.87-15.15), 50-59 (AOR: 4.65, 95% CI: 2.19-9.89), and ≥60 years (AOR: 12.67 95% CI: 5.46-29.39) had higher odds of central obesity compared to those below 30 years old. Likewise, Government/Non-governmental/Private employees (AOR: 4.68, 95% CI: 1.47-14.88), self-employed (AOR: 4.63, 95% CI: 1.62-13.24), and housewives (AOR: 5.21, 95% CI: 1.85-14.62) were more likely to be centrally obese compared to unemployed or students. Moreover, those individuals who skip breakfast were a 54% reduced risk of central obesity compared to their counterparts (**Table 5**).

**Table 5:** Factors associated with central obesity among adult **men** in administrative towns of Bale zone, Southeast Ethiopia, 2021.

| Variables | Abdominal obesity |  | COR (95% CI) | AOR<sup>a</sup> (95% CI) |
| --- | --- | --- | --- | --- |
|  | Yes | No |  |  |
|  | n (%) | n (%) |  |  |
| **Residential town** |  |  |  |  |
| Goba | 124 (43.4) | 162 (56.6) | 1.4 (1.0-1.8) | 1.10 (0.72-1.70) |
| Robe | 146 (36.0) | 260 (64.0) | **1** | **1** |
| **Sex** |  |  |  |  |
| Male | 40 (15.4) | 219 (84.6) | **1** | **1** |
| Female | 230 (53.1) | 203 (46.9) | 6.2 (4.2-9.1)* | 12.93 (6.74-24.79)** |
| **Age in years** |  |  |  |  |
| 18-29 | 56 (21.4) | 206 (78.6) | **1** | **1** |
| 30-39 | 59 (44.4) | 74 (55.6) | 2.9 (1.9-4.6)* | 2.80 (1.59-4.94)** |
| 40-49 | 61 (59.2) | 42 (40.8) | 5.3 (3.3-8.7)* | 7.66 (3.87-15.15)** |
| 50-59 | 37 (40.2) | 55 (59.8) | 2.5 (1.5-4.1)* | 4.65 (2.19-9.89)** |
| >=60 | 56 (56.0) | 44 (44.0) | 4.7 (2.9-7.7)* | 12.67 (5.46-29.39)** |
| **Marital status** |  |  |  |  |
| Never married | 19 (13.7) | 120 (86.3) | **1** | **1** |
| Married/cohabiting | 195 (42.1) | 268 (57.9) | 4.6 (2.7-7.7)* | 1.38 (0.63-3.05) |
| Divorced/separated/widowed | 56 (62.2) | 34 (37.8) | 10.4 (5.5-19.8)* | 1.05 (0.40-2.78) |
| **Educational status** |  |  |  |  |
| No formal education | 62 (63.3) | 36 (36.7) | 4.9 (2.9-8.5)* | 1.66 (0.70-3.90) |
| Primary education (1-8) | 100 (49.3) | 103 (50.7) | 2.8 (1.8-4.3)* | 1.38 (0.68-2.80) |
| Secondary education (9-12) | 64 (29.2) | 155 (70.8) | 1.2 (0.8-1.9) | 0.94 (0.48-1.86) |
| Diploma and above | 44 (25.7) | 127 (74.3) | **1** | **1** |
| **Occupational status** |  |  |  |  |
| House wives | 154 (61.1) | 98 (38.9) | 11.7 (6.0-22.4)* | 5.21 (1.85-14.62)** |
| Self employed | 57 (31.0) | 127 (69.0) | 3.3 (1.7-6.6)* | 4.63 (1.62-13.24)** |
| Government/private/NGO employed | 33 (29.2) | 80 (70.8) | 3.1 (1.5-6.3)* | 4.68 (1.47-14.88)** |
| Retired | 12 (33.3) | 24 (66.7) | 3.7 (1.5-9.3)* | 2.78 (0.73-10.48) |
| Student/unemployed | 12 (11.9) | 89 (88.1) | **1** | **1** |
| **Family size** |  |  |  |  |
| ≤ 2 | 28 (40.0) | 42 (60.0) | **1** | - |
| > 2 | 184 (37.9) | 301 (62.1) | 0.9 (0.5-1.5) | - |
| **Wealth index** |  |  |  |  |
| Low | 105 (41.3) | 173(58.7) | **1** | **1** |
| Medium | 60 (37.5) | 100 (62.5) | 0.9 (0.8-1.6) | 0.69 (0.40-1.17) |
| High | 105 (41.3) | 149 (58.7) | 1.2 (0.8-1.6) | 0.74 (0.45-1.22) |
| **Fruit and/or vegetable consumption on average per day** |  |  |  |  |
| Less than five servings per day | 128 (36.4) | 224 (63.6) | 0.8 (0.5-1.1) | 0.87 (0.57-1.32) |
| Five or more servings per day | 108 (43.0) | 143 (57.0) | **1** | **1** |
| **Avoid eating foods prepared outside of a home** |  |  |  |  |
| Yes | 245 (42.1) | 337 (57.9) | **1** | **1** |
| No | 21 (19.8) | 85 (80.2) | 0.3 (0.2-0.6)* | 1.59 (0.79-3.18) |
| **Skip breakfast** |  |  |  |  |
| No | 242 (41.0) | 348 (59.0) | **1** | **1** |
| Yes | 27 (27.0) | 73 (73.0) | 0.1 (0.3-0.9)* | 0.46 (0.23-0.90)** |
| **Level of physical activity** |  |  |  |  |
| Insufficient physical activity | 96 (49.5) | 98 (50.5) | 1.9 (1.3-2.6)* | 0.84 (0.55-1.30) |
| Sufficient physical activity | 166 (34.5) | 315 (65.5) | **1** | **1** |

**Notes:** **n**, frequency; **COR**, crude odds ratio; **AOR**, adjusted odds ratio; **CI**, confidence interval; <sup>a</sup>The model adjusted for: residence, sex, age, marital status, education status, occupation, fruit and vegetable consumption, avoiding eating foods prepared outside the home, skipping breakfast and level of physical activity; \*significant at $p < 0.05$ (crude); \*\*significant at $p < 0.05$ (adjusted).

## Discussion

The findings of this study reveal that the overall prevalence of central (abdominal) obesity was 39.0%. In comparison to men, women had a higher prevalence of central obesity (53.1% vs 15.4%). The figures reported in this study are comparable with studies conducted in Gondar and Dabat towns, Northwest Ethiopia (37.6% & 33.6%) (37, 43), but higher than the studies conducted in Nekemte town, West Ethiopia (28.4%) (35), Woldia town, Northeast Ethiopia (15.5%) (36), and Dilla town South Ethiopia (24.4%) (38). The possible variations of the obesity prevalence could be explained by the use of different cutoff values for waist circumferences (36) and the variation of age distributions between the studies participants (35, 38). Furthermore, variation in the gender make-up of the samples might be a possible reason for the differences, given such large differences in prevalence found in this study. Though there are slight variations in the prevalence rates across the regions, the magnitude of central obesity in the study appeared to be high. A recent systematic review and meta-analysis in Ethiopia reported that the prevalence of overweight and obesity is increasing especially in urban settings (44). The results of the present study confirm these findings and suggest that gender is a particularly important factor to examine when estimating overall prevalence rates in Ethiopian regions.

In this study, women had 13 times higher odds of central obesity compared to men. This finding is corroborated by similar studies in different parts of Ethiopia (35-39). Similarly, studies conducted by *Jaacks et al*., suggested that in countries with stage 1 obesity transition, the prevalence of obesity is higher among women compared to men (45). According to a study conducted by the Global Burden of Diseases report, the trend of obesity and overweight is persistently higher among women than men in developing countries (4). Further appropriately powered longitudinal studies within males and females in Ethiopia examining predictors of obesity/central obesity are warranted.

This study revealed that the odds of central obesity tend to increase as age increases. A longitudinal study conducted by *Baum & Ruhm* reported that body weight increases as age increases (46). The positive association between age and abdominal obesity is supported by several other studies conducted elsewhere including in Ethiopia, China, Russian Federation (35-37, 39, 47-50). This association might be explained partly by the gradual decline in physiologic activities and basal metabolic rate as age increases (51).

Employed adults and housewives had higher odds of central obesity compared to unemployed or students. This might be explained by the hypothesis that employed adults might have better access to foods than their counterparts. Previous studies in Ethiopia suggested that unemployed men are less likely to be obese compared to employed men (52). This finding contradicts a study conducted in North Glasgow, UK, which reported that unemployed men and women were less likely to be centrally obese compared to full-time workers (53). This contradiction needs to be addressed by conducting strong epidemiological studies exploring the role of employment status on the development of abdominal obesity in low-income countries. With regarding to findings of the higher odds of central obesity among housewives, it is plausible to hypothesis that staying at home exposes women to ease reach of food. As has stated elsewhere, frequency of meal intakes has been associated with an increase in total energy intake (54), hence increased opportunities to developing obesity.

Individuals who reported skipping their breakfast were 54% less likely to be centrally obese compared to their counterparts. This might be partly explained by the fact that frequency of meals is associated with an increase in total energy intake (54). However, this finding disagreed with a systematic review and meta-analysis conducted by *Ma* et al., which reported skipping breakfast results in a 31% increase in abdominal obesity (55). In this study, breakfast skipping behavior was measured through self-report and the reason for skipping breakfast was not collected. Moreover,meal frequency and portion size were not assessed, which could be a source of variation with existing evidence. Furthermore, this study was not topic-specific for the association between skipping breakfast and the risk of central obesity.

This study aimed to examine predictors of central (abdominal) obesity in a region with no prior evidence on the distribution and associated risk factors. Although the study used primary data on physical measurements like waist and hip circumferences and the WHO-STEPS wise tool for non-communicable disease risk factors surveillance with trained data collectors and supervisors, the findings must be interpreted in light of the following limitations. First, due to the cross-sectional nature of the study, a cause-effect relationship cannot be established between the risk factors and obesity. Secondly, the study sample was too small for sex and age-specific reporting of the prevalence and risk factors. Differences in prevalence and risk factors by gender suggest the need for gender-specific stratification in larger, appropriately powered longitudinal studies. Lastly, data regarding dietary habits, physical activity, alcohol, and tobacco use were collected through self-reported behavior questionnaires, which might be affected by the recall and social desirability bias. Future studies could use more objective measurements, including third part reporting and wearables to assess these behaviors.

## Conclusions

The burden of abdominal obesity is high in southeast Ethiopia, especially in women, with one out of every two women and three out of every 10 men found to be centrally (abdominally) obese. Prevention strategies that account for identified modifiable risk factors are needed to curb the increasing burden of central obesity and its public health impact. Further studies of gender-specific risk factors are warranted.

## Data Availability

All relevant data are within the manuscript and its Supporting Information files.

## Acknowledgments

We thank Madda Walabu University for the financial support to conduct this study. We are also grateful to all data collectors, Goba and Robe Health Office and Health extension workers, kebele and Got administrators for their facilitation and cooperation to smoothly conduct this study.

## Supporting information

**Figure 1:**
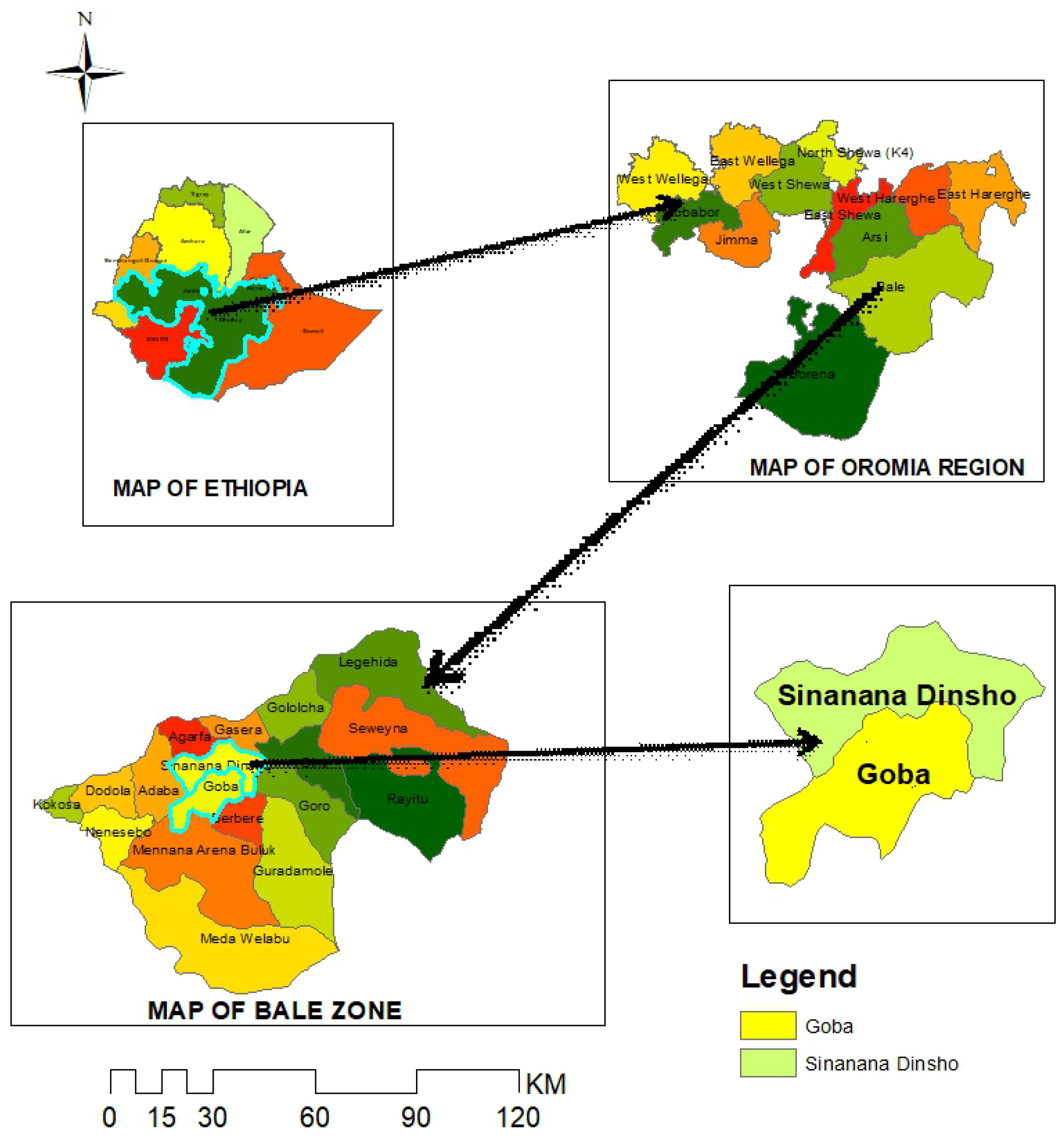
Map of Goba and Robe town, Southeast Ethiopia

## Notes

### Competing Interest Statement

The authors have declared no competing interest.

### Funding Statement

The funders had no role in study design, data collection and analysis, decision to publish, or preparation of the manuscript.

